# An Automated CT-Derived Marker of Renal Tumor Complexity: The CLARITY Score

**DOI:** 10.64898/2026.05.08.26352647

**Authors:** Rishi Jonnalagadda, Sahil Patel, Haya T. Abusafieh, Rikhil Seshadri, Daniel Jevnikar, Salim Younis, Abdulrahman Al-Bayati, Nicolas Saputro, Jacob M. Knorr, Betty Wang, Michal Ozery-Flato, Michal Rosen-Zvi, Robert Abouassaly, Erick Remer, Nicholas E. Heller, Christopher J. Weight

## Abstract

**Background and Objective:** Surgical complexity for renal tumors has traditionally been assessed using manual nephrometry scores, which require unreimbursed physician effort and are subject to interobserver variability. This study introduces an objective, fully automated alternative derived from decades of experience at a large academic center.

**Methods:** We trained a CT classification model to predict whether a patient would ultimately undergo Partial or Radical Nephrectomy (PN or RN). We hypothesized that the model’s confidence in RN (termed the CLARITY score) would serve as a surrogate for the difficulty of nephron-sparing approaches and thus for tumor complexity. This hypothesis was tested using multivariate logistic regression for failure to achieve trifecta, estimated blood loss (EBL) ≥ 500 mL, and length of stay ≥ 3 d. CLARITY was compared with tumor size and R.E.N.A.L. score. External validation in a geographically distinct cohort was performed.

**Key Findings and Limitations:** For predicting RN, CLARITY achieved an AUROC of 0.899 internally and 0.898 externally. In the external PN subgroup, it outperformed tumor size and R.E.N.A.L. score in predicting failure to achieve trifecta (AUROC 0.613), EBL ≥ 500 mL (0.727), and length of stay ≥ 3 d (0.673). In multivariable analysis, CLARITY remained associated with each outcome, whereas R.E.N.A.L. and size were not. This study is limited by its retrospective design.

**Conclusions and Clinical Implications:** CLARITY is an automated CT-derived marker that quantifies renal tumor complexity more effectively than tumor size and R.E.N.A.L. score and may support scalable, objective preoperative complexity assessment. To support reproducibility and external validation, we have released a public inference pipeline and web-based DICOM upload portal for research use.

## Introduction

Partial nephrectomy (PN) is preferred for many localized renal masses because it preserves postoperative renal function. However, the technical difficulty of nephron-sparing surgery varies substantially across cases, and more difficult PN is associated with greater bleeding, prolonged ischemia, complications, and longer hospitalization.^1^ Accurate preoperative assessment of nephron-sparing complexity is therefore important for counseling, case selection, referral, and operative planning.

Current preoperative complexity assessment relies largely on tumor size and manual nephrometry systems such as R.E.N.A.L. and PADUA.^2,3^ These tools provide a structured framework for anatomic characterization and have been widely studied in relation to peri-operative outcomes. However, their predictive performance is variable across settings, and manual scoring requires unreimbursed time, is subject to interobserver variability, and is not consistently incorporated into routine workflows.^4,5,6^ There remains a need for an objective and scalable imaging-based marker of renal tumor complexity.

Preoperative contrast-enhanced CT contains rich anatomic information over and above what is captured by ordinal nephrometry scores. Prior work has shown that nephrometry components and composite scores can be automated from CT.^7,8^ However, most such efforts have focused on reproducing existing scoring systems rather than empirically deriving new imaging markers that more directly reflect operative complexity.

We evaluated a CT-based deep learning model trained to predict whether patients would ultimately undergo radical nephrectomy (RN) rather than partial nephrectomy (PN). We then repurposed the model’s predicted probability of RN as a marker of nephron-sparing complexity, reasoning that imaging features associated with a surgeon’s choice of RN likely capture the characteristics that make a tumor more complex. We term this probability the “CLeveland Automated Renal tumor complexITY” (or “CLARITY”) score.

The aim of this study was to determine (a) whether CLARITY associates with adverse outcomes from partial nephrectomy, as we would expect from a good measure of complexity, and (b) whether this score generalizes to another institution where surgical decision making is carried out by an entirely different group of surgeons, or for patients from different geographical regions. Importantly, we directly compare this new measure’s performance against the impressive benchmarks set by tumor size and traditional nephrometry scores.

## Materials (Patients) and Methods

This retrospective study was approved by our institution’s IRB (study ID 23-1158); the requirement for informed consent was waived due to retrospective design and minimal risk. Using CPT codes, we identified 9096 patients who underwent nephrectomy for a renal tumor between October 2009 and July 2024. Patients without available preoperative contrast-enhanced CT imaging or missing endpoint data were excluded, leaving a cohort of 1628 nephrectomy patients with complete data, detailed by Table 1.

**Table 1:** Baseline demographic and clinical characteristics of the internal cohort stratified by surgical approach. Values are presented as median [interquartile range] or n (%). PN = partial nephrectomy; RN = radical nephrectomy.

| Characteristic | Overall (N=1628) | PN (n=1075) | RN (n=553) | *p* value |
| --- | --- | --- | --- | --- |
| Age at surgery, yr | 64.7 [55.3–71.7] | 63.9 [54.6–70.7] | 65.6 [57.0–73.3] | < 0.001 |
| **Sex** |  |  |  | 0.147 |
| Male | 1004 (61.7%) | 649 (60.4%) | 355 (64.2%) |  |
| Female | 624 (38.3%) | 426 (39.6%) | 198 (35.8%) |  |
| **Race** |  |  |  | 0.326 |
| White | 1527 (93.8%) | 1008 (93.8%) | 519 (93.9%) |  |
| Black | 80 (4.9%) | 57 (5.3%) | 23 (4.2%) |  |
| Asian | 5 (0.3%) | 3 (0.3%) | 2 (0.4%) |  |
| American Indian/<br>Alaska Native | 5 (0.3%) | 2 (0.2%) | 3 (0.5%) |  |
| Other/Multiracial | 11 (0.7%) | 5 (0.5%) | 6 (1.1%) |  |
| **Ethnicity** |  |  |  | 0.055 |
| Not Hispanic/Latino | 1531 (94.0%) | 1003 (93.3%) | 528 (95.5%) |  |
| Hispanic/Latino | 68 (4.2%) | 54 (5.0%) | 14 (2.5%) |  |
| Unknown | 29 (1.8%) | 18 (1.7%) | 11 (2.0%) |  |
| BMI, kg/m<sup>2</sup> | 30.0 [26.1–34.2] | 30.2 [26.5–34.5] | 29.2 [25.7–33.9] | 0.010 |
| Radiographic tumor size, cm | 4.0 [2.7–6.3] | 3.2 [2.4–4.5] | 6.7 [5.0–9.1] | < 0.001 |
| Preoperative metastasis | 65 (4.0%) | 16 (1.5%) | 49 (9.0%) | < 0.001 |
| Preoperative<br>lymphadenopathy | 98 (6.0%) | 39 (3.6%) | 59 (10.7%) | < 0.001 |
| **Surgical approach** |  |  |  | < 0.001 |
| Robotic | 971 (59.6%) | 756 (70.3%) | 215 (38.9%) |  |
| Open | 422 (25.9%) | 259 (24.1%) | 163 (29.5%) |  |
| Laparoscopic | 235 (14.4%) | 60 (5.6%) | 175 (31.6%) |  |
| **Laterality** |  |  |  | 0.161 |
| Right | 856 (52.6%) | 547 (50.9%) | 309 (55.9%) |  |
| Left | 769 (47.2%) | 526 (48.9%) | 243 (43.9%) |  |
| Bilateral | 3 (0.2%) | 2 (0.2%) | 1 (0.2%) |  |

External validation was performed in an independent, publicly available nephrectomy cohort from the Kidney Tumor Segmentation (KiTS) dataset,^9^ which includes contrast-enhanced CT imaging and clinical data from patients undergoing partial or radical nephrectomy at institutions independent of the development cohort. Patients with available preoperative imaging and complete perioperative outcome data were included, yielding an external validation cohort of *n* = 452. This cohort allowed evaluation of CLARITY across different imaging acquisition, surgical decision-making, and perioperative care patterns. R.E.N.A.L. nephrometry scores in this cohort were manually assigned by three independent reviewers and consolidated by averaging.

We used a multiple instance learning framework to train a CT-based deep learning model to distinguish partial from radical nephrectomy. Cases in which the case was converted from partial to radical during the procedure were counted as radicals. Technical details regarding preprocessing, sampling, architecture, and training are provided in the Supplementary Material section S1. The model produces a continuous value representing its confidence that the patient will undergo radical nephrectomy, ranging from 0 to 1, which we call the “CLARITY” score. Thus, higher CLARITY scores indicate imaging patterns more strongly associated with a surgeon choosing to perform radical nephrectomy and presumably with less favorable anatomy for nephron-sparing surgery.

In the external validation cohort, CLARITY was evaluated in two complementary ways. First, we assessed its discrimination, calibration, and decision-curve characteristics for prediction of PN versus RN. Second, we evaluated it as a preoperative imaging marker of partial nephrectomy complexity by comparing its association with perioperative endpoints against tumor size and consensus R.E.N.A.L. score.

Because preoperative complexity assessment is most relevant when nephron-sparing surgery is attempted, primary clinical analyses are restricted to the partial nephrectomy subgroup. Endpoints were failure to achieve trifecta, estimated blood loss (EBL) ≥ 500 mL, and length of stay ≥ 3 d. Failure to achieve trifecta was defined as positive surgical margin, warm ischemia time *>* 25 min, or any postoperative complication.^10^

To improve interpretability of the continuous CLARITY score, we performed a stratification into low-, intermediate-, and high-complexity bands. These strata were derived using an unsupervised three-component Gaussian mixture model fit only to CLARITY scores in the internal cohort.^11^ Component boundaries were defined as the score values where the posterior probabilities of adjacent mixture components were equal. The resulting thresholds were then applied without refitting to the external validation cohort. The mixture model used CLARITY scores only and did not use surgical approach or perioperative outcome labels. Outcome rates were summarized across CLARITY bands in the external partial nephrectomy subgroup, and radical nephrectomy prevalence was summarized across bands in the full external cohort.

Additional supportive analyses compared CLARITY with four preoperative complexity markers: AI-derived continuous R.E.N.A.L. score, manually assigned discrete R.E.N.A.L. score, PADUA score, and tumor size. Manually assigned PADUA scores were included for exploratory analyses. The AI-derived continuous R.E.N.A.L. score was derived from model-predicted probabilities for individual R.E.N.A.L. nephrometry components, as described previously.^7,8^

For binary endpoints, marker discrimination was assessed using the area under the receiver operating characteristic curve (AUROC). Differences in AUROC were compared using DeLong’s test for correlated ROC curves.^12^ Continuous associations were assessed using Spearman correlation. Multivariable logistic regression was performed for failure to achieve trifecta, EBL ≥ 500 mL, and length of stay ≥ 3 d to assess whether the CLARITY score remained associated with outcome after adjustment for R.E.N.A.L. score and tumor size. All tests were two-sided, with statistical significance defined as *p <* 0.05.

## Results

The internal cohort included 1,628 patients for model development and testing. Baseline demographic and clinical characteristics of the internal cohort are shown in Table 1. For discrimination of RN versus PN, CLARITY achieved an AUROC of 0.899, outperforming tumor size (0.862; DeLong *p* = 3.7 *×* 10^−6^) and R.E.N.A.L. score (0.804; DeLong *p* = 1.2 *×* 10^−17^) (Figure 1). In the subset of tumors 4–9 cm where the PN vs. RN decision is most challenging (Supplementary Figure S6), CLARITY achieved an AUROC of 0.834 versus 0.732 for tumor size and 0.725 for R.E.N.A.L. score (both *p <* 0.001). These findings support that the score captures imaging information relevant to nephron-sparing favorability beyond conventional preoperative markers. In the independent external validation cohort (*n* = 452), CLARITY showed similarly strong discrimination between RN and PN, with an AUROC of 0.898. The Brier score was 0.126 in the external cohort and 0.118 in the internal cohort, meaning the model was well-calibrated in both cohorts (Figure 1; Supplementary Figure S2).

**Figure 1:**
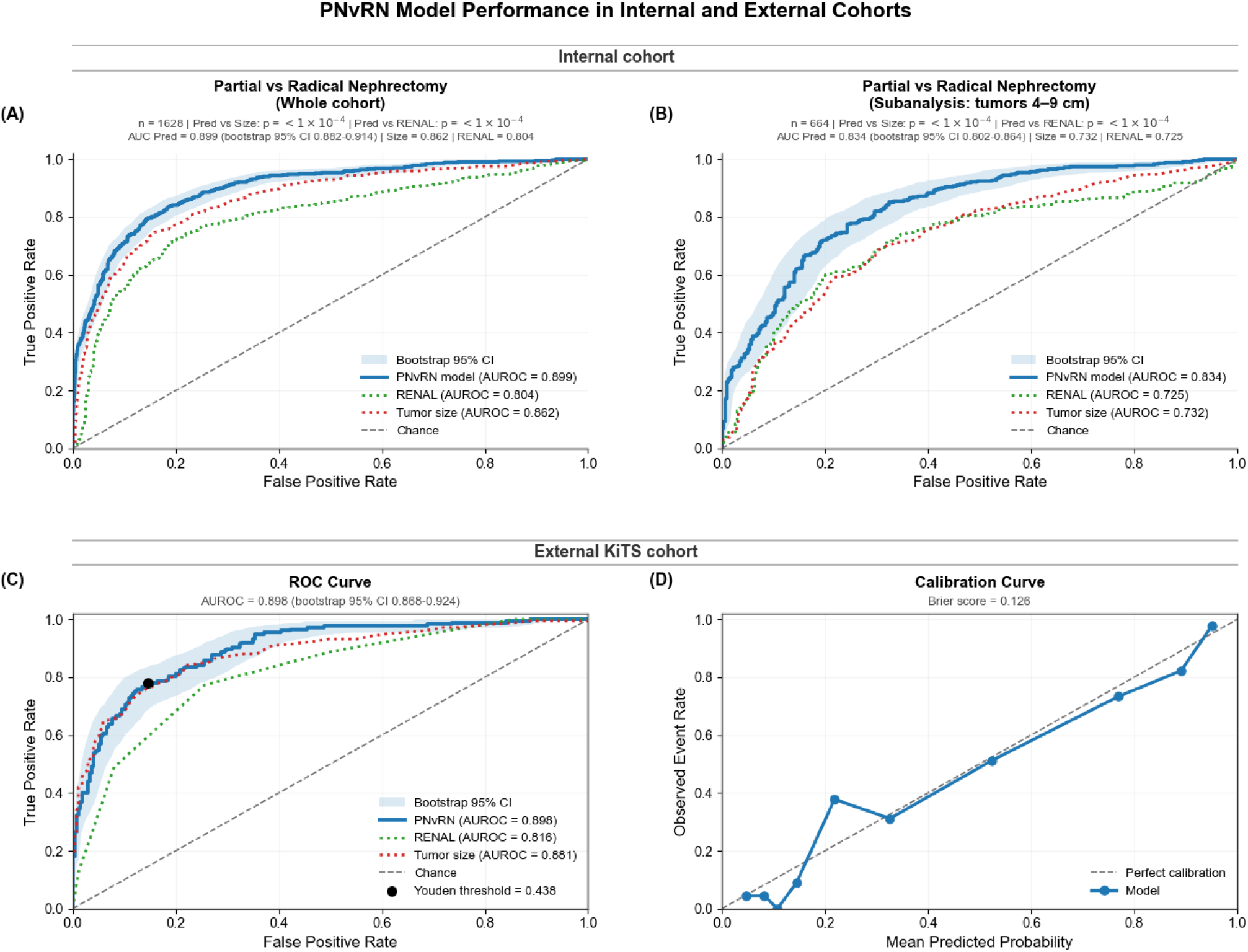
Classification of partial versus radical nephrectomy in the internal and external validation cohorts. (A) Receiver operating characteristic (ROC) curves for prediction of partial versus radical nephrectomy in the full internal cohort. (B) ROC curves for prediction of partial versus radical nephrectomy in the 4–9 cm subgroup. (C) ROC curve for prediction of partial versus radical nephrectomy in the full external cohort. (D) Calibration curve for the external cohort. PN = partial nephrectomy; RN = radical nephrectomy; ROC = receiver operating characteristic.

For failure to achieve trifecta, CLARITY achieved an AUROC of 0.613, compared with 0.510 for R.E.N.A.L. score and 0.525 for tumor size. The strongest domain-level signal was observed for major blood loss, for which CLARITY achieved an AUROC of 0.727, compared with 0.620 for R.E.N.A.L. score and 0.657 for tumor size. For length of stay ≥ 3 d, CLARITY also showed the strongest discrimination (AUROC 0.673 vs 0.604 for R.E.N.A.L. score and 0.615 for tumor size). Improvement over R.E.N.A.L. score was significant for all three endpoints (Table 2, Figure 2B).

**Table 2:** AUROCs of CLARITY, R.E.N.A.L. score, and tumor size across the main partial nephrectomy complexity endpoints in the external validation cohort.

| Endpoint | CLARITY<br>(95% CI) | R.E.N.A.L.<br>(95% CI) | Tumor size<br>(95% CI) | *p* vs R.E.N.A.L. | *p* vs size |
| --- | --- | --- | --- | --- | --- |
| Failure to achieve trifecta | 0.613<br>(0.545–0.683) | 0.510<br>(0.444–0.580) | 0.525<br>(0.451–0.599) | 0.0030 | 0.0062 |
| EBL ≥ 500 mL | 0.727<br>(0.649–0.796) | 0.620<br>(0.528–0.705) | 0.657<br>(0.559–0.745) | 0.0026 | 0.1055 |
| LOS ≥ 3 d | 0.673<br>(0.606–0.735) | 0.604<br>(0.533–0.670) | 0.615<br>(0.546–0.681) | 0.0417 | 0.0674 |

AUROC = area under the receiver operating characteristic curve; CI = confidence interval; EBL = estimated blood loss; LOS = length of stay.

**Figure 2:**
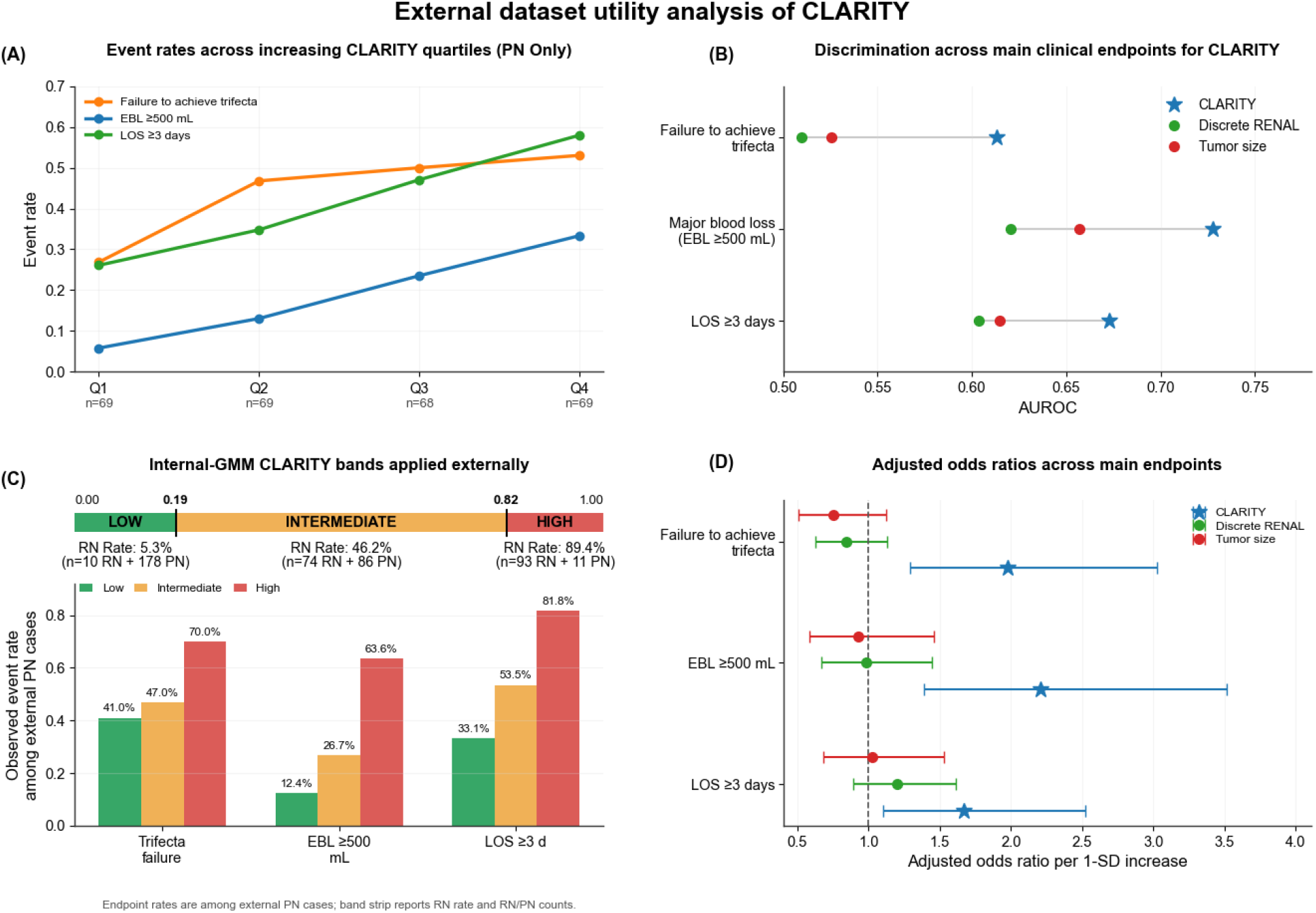
External validation of the CLARITY score as a marker of partial nephrectomy complexity. Failure to achieve trifecta, estimated blood loss (EBL) ≥ 500 mL, and length of stay (LOS) ≥ 3 d were examined. (A) Event rates across CLARITY score quartiles among patients undergoing partial nephrectomy. (B) Area under the receiver operating characteristic curve (AUROC) comparison across the same endpoints. (C) Internally derived score bands and observed external partial nephrectomy outcome rates across bands. (D) Adjusted odds ratios across endpoints and predictors. AUROC = area under the receiver operating characteristic curve; EBL = estimated blood loss; LOS = length of stay; OR = odds ratio; GMM = Gaussian mixture model.

The clinical gradient across CLARITY scores further supported its relevance as a complexity marker. From the lowest to highest CLARITY score quartile, the rate of failure to achieve trifecta increased from 26.9% to 53.0%, EBL ≥ 500 mL from 5.8% to 33.3%, and length of stay ≥ 3 d from 26.1% to 58.0% (Figure 2A). Thus, higher CLARITY scores identified a progressively higher-burden perioperative phenotype among patients undergoing PN.

Applying the internally derived CLARITY bands to the external validation cohort showed a clear gradient in surgical approach and perioperative burden. In the full external cohort, radical nephrectomy rates were 5.3%, 46.2%, and 89.4% across the low-, intermediate-, and high-complexity bands, respectively. Among patients undergoing partial nephrectomy, EBL ≥ 500 mL increased from 12.4% to 26.7% and 63.6%, and LOS ≥ 3 d increased from 33.1% to 53.5% and 81.8%. Failure to achieve trifecta was also most frequent in the high-complexity band (41.0%, 47.0%, and 70.0%; Figure 2C).

In multivariable analysis adjusting for tumor size and R.E.N.A.L. score, CLARITY remained independently associated with failure to achieve trifecta (odds ratio [OR] 1.98 per standard deviation increase, 95% confidence interval [CI] 1.29–3.03; *p* = 0.0017), EBL ≥ 500 mL (OR 2.21, 95% CI 1.39–3.51; *p* = 0.0008), and length of stay ≥ 3 d (OR 1.67, 95% CI 1.10–2.52; *p* = 0.015) (Figure 2D).

Supportive exploratory analyses compared CLARITY with four additional preoperative complexity markers: AI-derived continuous R.E.N.A.L. score, manually assigned discrete R.E.N.A.L. score, PADUA score, and tumor size. Across the evaluated endpoints, CLARITY had the highest AUROC among the five predictors for failure to achieve trifecta, EBL ≥ 500 mL, LOS ≥ 3 d, positive margin, major complications, and any complication. CLARITY also had the strongest Spearman correlation with length of hospitalization. Full exploratory results are shown in Figure 3.

**Figure 3:**
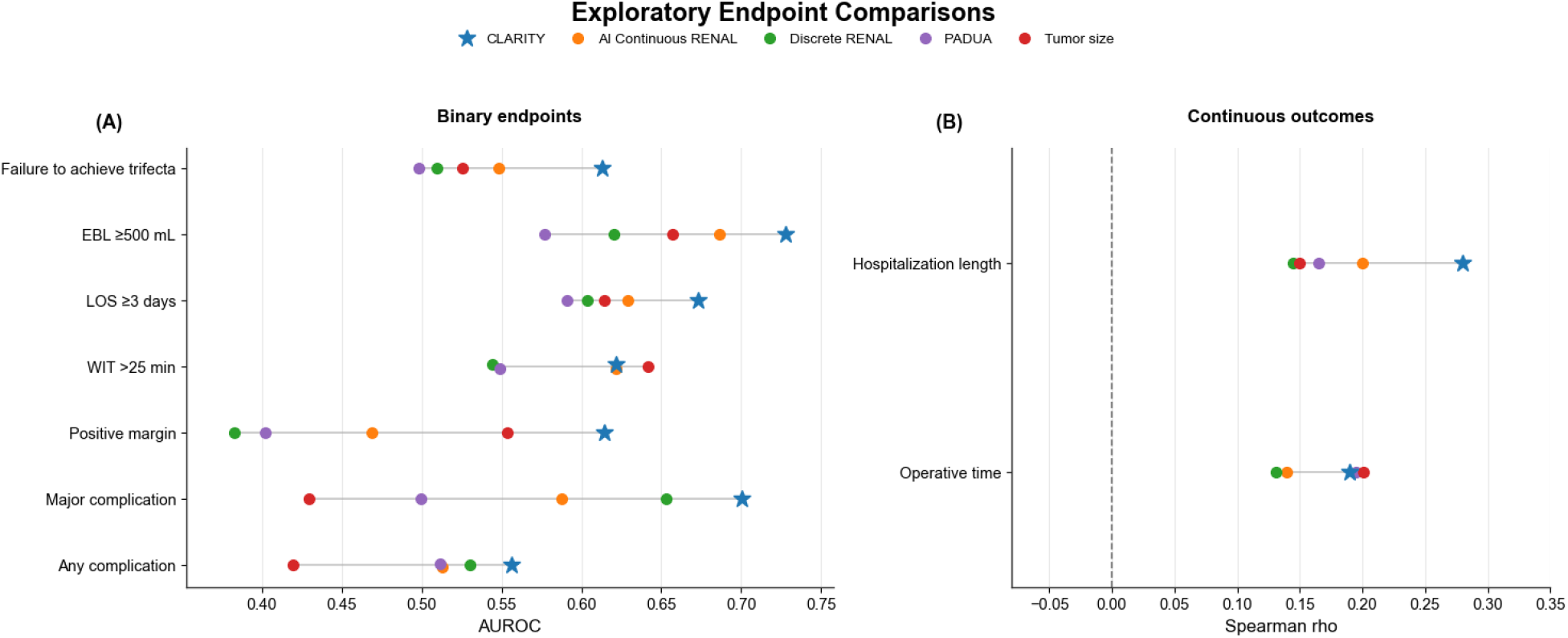
Exploratory comparison of five preoperative complexity markers across additional perioperative endpoints in the external validation cohort: CLARITY, AI-derived continuous R.E.N.A.L. score, manually assigned discrete R.E.N.A.L. score, PADUA score, and tumor size.

## Discussion

Our findings suggest that a CT-derived deep learning score can function as an automated imaging marker of partial nephrectomy complexity. Although CLARITY was trained to distinguish partial from radical nephrectomy, its clinically meaningful value emerges within the partial nephrectomy population itself: on external evaluation, higher CLARITY scores consistently identified patients with greater perioperative burden, including higher rates of trifecta failure, major blood loss, and prolonged hospitalization.

The principal clinical question was whether CLARITY could identify more difficult partial nephrectomies among patients in whom nephron-sparing surgery was attempted. In the external partial nephrectomy cohort, CLARITY demonstrated the strongest discrimination across all three prespecified complexity endpoints—failure to achieve trifecta, EBL ≥ 500 mL, and length of stay ≥ 3 d—when compared with tumor size and R.E.N.A.L. score. In addition to these discrimination results, the observed quartile-based gradients in perioperative outcomes support its clinical relevance, with progressively higher complication burden across increasing CLARITY strata. The internally derived low-, intermediate-, and high-complexity bands provide an initial reporting framework for interpreting the continuous score, although these exploratory cut points require prospective validation before use as clinical decision thresholds. Systematic analysis correlating attention-derived spatial explanations with established nephrometry components has not been performed and represents an important direction for future work. Representative CT examples spanning the score range further illustrate the underlying radiographic patterns captured by the model, and align well with expert intuition (Figure 4).

**Figure 4:**
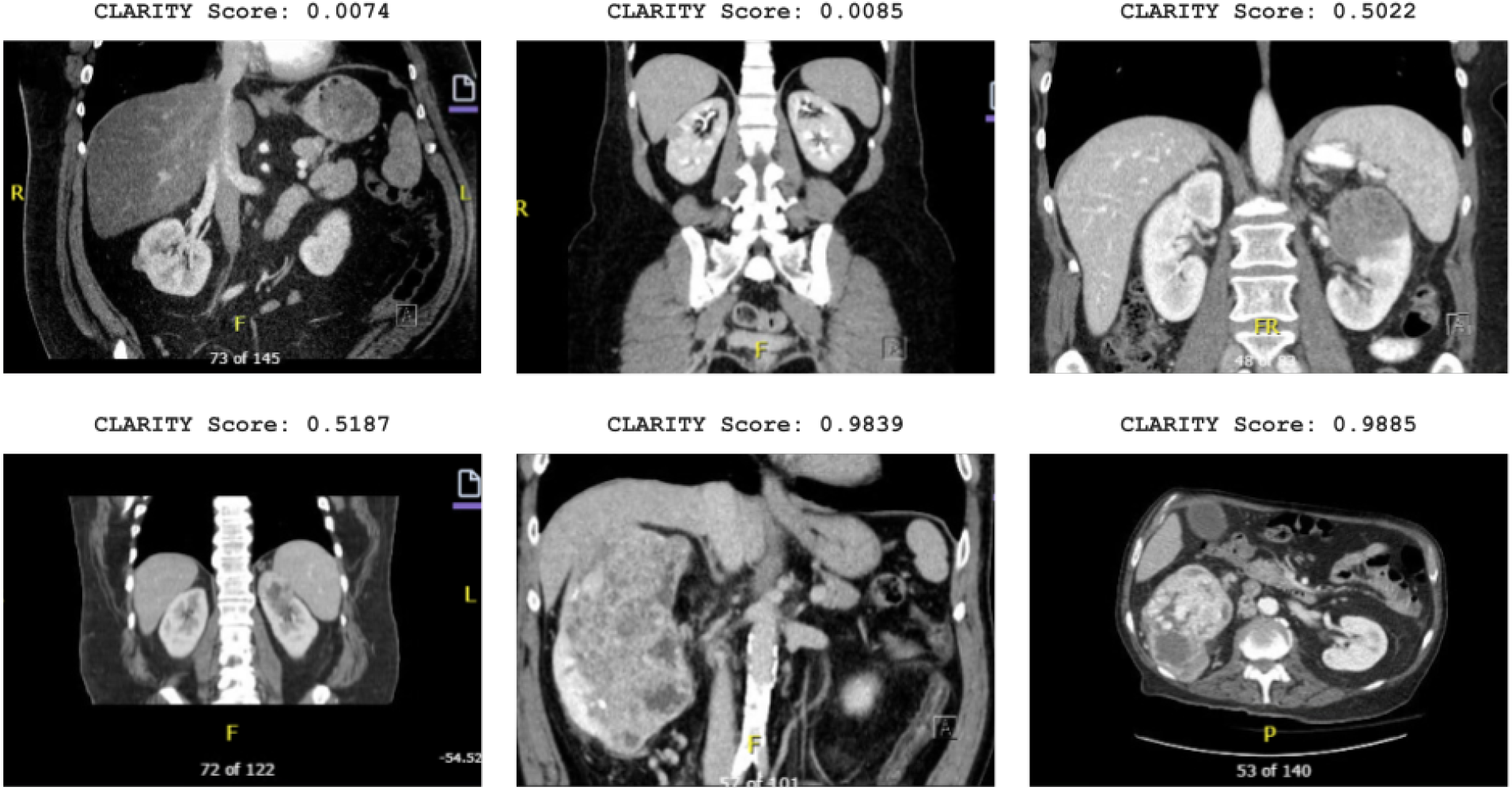
Representative preoperative CT images spanning a range of CLARITY scores in the internal dataset. Included are the two CT scans with the lowest CLARITY scores, the two scores closest to 0.5, and the two highest scores. Lower scores correspond to imaging patterns more consistent with less complex partial nephrectomy, whereas higher scores correspond to imaging patterns more strongly associated with greater operative complexity. CT = computed tomography.

The close agreement between internal and external performance supports portability of the learned representation and justifies its application as an objective imaging marker of renal tumor complexity rather than an idiosyncratic classifier of surgical approach at a single institution. This is particularly relevant given that many prior imaging-based artificial intelligence models remain limited to internal validation, especially for tasks influenced by surgeon preference. Here, both surgical approach discrimination and complexity associations were preserved across institutions, strengthening confidence in the generalizability of the approach.

Manual nephrometry systems remain valuable for structured anatomic description and standardized reporting, but they require physician effort, are subject to interobserver variability, and are not consistently incorporated into routine workflows. CLARITY approaches the same problem from a different perspective: rather than assigning discrete anatomic components, it derives a continuous score directly from imaging and appears to capture composite features associated with operative difficulty. The attention-based pooling mechanism intrinsic to the model architecture assigns importance weights to a sample of individual image patches, providing a degree of spatial interpretability consistent with inherently interpretable AI design, an increasingly valued property in clinical AI.^13,14^ It is therefore best viewed not as a replacement for surgical judgment or nephrometry, but as an automated and scalable complement that may facilitate reproducible complexity assessment, particularly in large-scale studies or workflow-integrated decision support. To facilitate reproducibility and external testing, we have also released a public inference pipeline and web-based DICOM upload portal that allows users to obtain CLARITY scores from preoperative imaging. Prospective studies will be needed to evaluate how such implementation affects clinical workflow, counseling, and decision-making.

Among the evaluated endpoints, the strongest signal was observed for major blood loss. This is clinically plausible, as intraoperative bleeding during partial nephrectomy is closely tied to tumor centrality, hilar proximity, and the technical complexity of resection and reconstruction—features that are inherently encoded in preoperative imaging. The consistent performance across endpoints, combined with the observed gradients in outcome risk, supports the interpretation of CLARITY as a meaningful representation of operative complexity.

Several limitations should be considered. First, the derivation target of partial versus radical nephrectomy reflects surgical decision-making rather than a purely anatomic ground truth and may therefore incorporate practice patterns in addition to tumor complexity. Second, the retrospective design introduces potential selection bias. Third, downstream analyses were structured as marker comparisons rather than endpoint-specific predictive models, and observed AUROCs for some endpoints were modest. Accordingly, CLARITY should be interpreted as a complexity marker rather than a standalone predictor of perioperative outcomes. Fourth, the low-, intermediate-, and high-complexity CLARITY bands were exploratory, and the high-complexity partial nephrectomy subgroup in the external cohort was small; these thresholds should therefore be viewed as interpretive reporting categories rather than validated clinical decision thresholds, which would require further prospective validation. Fifth, the internal cohort is predominantly White (93.8%), and racial and ethnic diversity in both cohorts is limited. Whether CLARITY performs equitably across more diverse populations, in whom tumor biology, access to nephron-sparing surgery, and baseline comorbidity profiles may differ, remains to be established, and validation in more demographically representative cohorts is warranted. Finally, although the external cohort provides independent and publicly reproducible validation, it was not designed specifically for prospective complexity prediction, and prospective evaluation within clinical workflows will be necessary to determine real-world utility.

Taken together, these findings suggest that preoperative CT can be leveraged not only to automate existing nephrometry systems, but also to derive a more direct and scalable imaging-based measure of nephron-sparing complexity. Further multicenter validation, prospective implementation studies, and integration with clinical variables represent important next steps toward clinical translation.

## Conclusions

CLARITY is a fully automated, externally evaluated CT-derived imaging marker of partial nephrectomy complexity. In an independent external cohort, higher CLARITY scores were associated with adverse perioperative outcomes among patients undergoing partial nephrectomy and outperformed manual nephrometry for key complexity endpoints. Together with its public inference pipeline, CLARITY provides a scalable framework for reproducible preoperative complexity assessment and future external validation.

## Data Availability

To support reproducibility and independent validation, CLARITY score predictions for the KiTS external validation cohort are publicly available as a JSON file at the project repository. Public inference code, model weights, and a containerized implementation are available at the CLARITY inference pipeline repository. The released pipeline accepts DICOM imaging input, performs preprocessing and segmentation, and returns a CLARITY score for each case.
A web-based CLARITY upload portal is also available at clarity.aim-hi-lab.org/. Users may upload a preoperative kidney tumor DICOM case and provide an email address to receive a CLARITY score after processing. The portal is intended to support reproducibility, external testing, and prospective workflow evaluation.
Because external KiTS validation in this study was performed using the segmentations and NIfTI images provided by the KiTS dataset rather than pipeline-generated segmentations, reproduced results may differ due to variation in segmentation quality and preprocessing. The data from the internal dataset will not be provided as it includes protected health information.

https://github.com/AIM-HI-Lab/axis-inference-pipeline/blob/main/paper_data/inference_aggregated.json

https://github.com/AIM-HI-Lab/axis-inference-pipeline

https://clarity.aim-hi-lab.org

## Open Science Statement

To support reproducibility and independent validation, CLARITY score predictions for the KiTS external validation cohort are publicly available as a JSON file at the project repository. Public inference code, model weights, and a containerized implementation are available at the CLARITY inference pipeline repository. The released pipeline accepts DICOM imaging input, performs preprocessing and segmentation, and returns a CLARITY score for each case.

A web-based CLARITY upload portal is also available at clarity.aim-hi-lab.org. The portal is intended to support reproducibility, external testing, and prospective workflow evaluation using de-identified preoperative imaging. The released model and portal are intended for research and validation use and should not be used for clinical decision-making.

Because external KiTS validation in this study was performed using the segmentations and NIfTI images provided by the KiTS dataset rather than pipeline-generated segmentations, reproduced results may differ due to variation in segmentation quality and preprocessing. The data from the internal dataset will not be provided as it includes protected health information.

## Ethics Approval

This retrospective study was approved by the Cleveland Clinic Institutional Review Board (study ID 23-1158). The requirement for informed consent was waived because of the retrospective design and minimal risk.

## CRediT Author Statement

Rishi Jonnalagadda: Conceptualization; Data curation; Formal analysis; Investigation; Methodology; Software; Validation; Visualization; Writing – original draft; Writing – review & editing.

Sahil Patel: Conceptualization; Investigation; Methodology; Validation; Writing – review & editing.

Haya T. Abusafieh: Data curation; Investigation; Writing – review & editing.

Rikhil Seshadri: Data curation; Investigation; Software; Writing – review & editing.

Daniel Jevnikar: Data curation; Investigation; Writing – review & editing.

Salim Younis: Data curation; Investigation; Writing – review & editing.

Abdulrahman Al-Bayati: Data curation; Investigation; Writing – review & editing.

Nicolas Saputro: Data curation; Investigation; Validation; Writing – review & editing.

Jacob M. Knorr: Data curation; Investigation; Validation; Writing – review & editing.

Betty Wang: Data curation; Investigation; Validation; Writing – review & editing.

Michal Ozery-Flato: Resources; Software.

Michal Rosen-Zvi: Methodology; Resources; Supervision; Writing – review & editing.

Robert Abouassaly: Resources; Supervision; Writing – review & editing.

Erick Remer: Resources; Writing – review & editing.

Nicholas E. Heller: Conceptualization; Methodology; Resources; Software; Supervision; Validation; Writing – review & editing.

Christopher J. Weight: Conceptualization; Funding acquisition; Project administration; Resources; Supervision; Writing – review & editing.

