## Supplemental Material for "An Automated CT-Derived Marker of Renal Tumor Complexity: The CLARITY Score"

This supplementary material accompanies the medRxiv preprint titled "An Automated CT-Derived Marker of Renal Tumor Complexity: The CLARITY Score."

### Supplementary Methods and Figures

#### S1. Training Procedure

##### Imaging Processing

Preoperative CT scans were retrieved in DICOM format and converted to a standardized orientation. All scans were resampled and intensity normalized prior to model input, with an HU range of $[-128, 256]$. Tumor segmentations were obtained using the methods of the top-scoring teams from the most recent KiTS challenge.[1]

##### Training Procedure

From each CT volume, two-dimensional image slices were extracted in the axial plane. Axial slices were selected because of their higher and more uniform native spatial resolution relative to reconstructed coronal or sagittal views. Image patches centered on the tumor and surrounding renal anatomy were sampled from each scan to create sets of image instances corresponding to each patient, but slices were assigned to the train and test sets at the patient level to avoid data leakage.

These sampled image patches collectively formed a bag of instances representing each patient case, enabling multiple instance learning training.

A patient-level held-out test set (20%) was separated before model training to avoid leakage. Within the remaining training cohort, five-fold cross-validation was used for model selection. The best model from each fold, as measured on that fold's validation set, was then run on the testing set and averaged to obtain the final results (Figure S3). A different 20% patient subset was then used as the test set, and the same process was repeated until all patients had been included in a held-out test set. Across the repeated held-out test partitions, a total of 25 fold-specific models contributed to the aggregated internal performance estimates.

During training, each patient case was represented by a set of sampled axial image patches forming a bag of instances. Data augmentation techniques, including random flips and rotations, were applied during training to improve generalization.

Optimization was performed using the Adam optimizer with an initial learning rate of $2 \times 10^{-4}$. We used a ReduceLROnPlateau schedule with a plateau patience of 4 epochs, plateau factor of 0.2, and a minimum learning rate of $1 \times 10^{-6}$. Training was performed for up to 100 epochs with early stopping after 10 validation epochs without improvement. Binary classification tasks were trained using binary cross-entropy loss. Models were trained on a single NVIDIA V100.

##### Model Architecture

We used the ResNet-18 architecture[2] pretrained on ImageNet and fine-tuned on our cohort of CT images. In pilot experiments, we obtained similar results using ResNet-34 and ResNet-50 architectures, so we selected the lighter-weight and faster ResNet-18 model.

A multiple instance learning framework was used to aggregate image embedding information across multiple patches belonging to the same patient. Each image patch was independently processed by the same shared CNN encoder to produce a feature representation. These instance-level features were then aggregated using an attention-based pooling mechanism,[3] which learns to assign importance weights to individual image patches. The aggregated feature representation was passed to a fully connected classification head to generate patient-level predictions for each endpoint (Figure S4).

#### S2. Exploratory Comparison with Additional Preoperative Complexity Markers

In addition to the primary comparisons emphasized in the main text, we performed supportive exploratory analyses comparing five preoperative complexity markers: CLARITY, AI-derived continuous R.E.N.A.L. score, manually assigned discrete R.E.N.A.L. score, PADUA score, and tumor size. The AI-derived continuous R.E.N.A.L. score was derived from model-predicted probabilities for the individual R.E.N.A.L. nephrometry components, as described previously.[4,5]

These exploratory analyses were intended to assess whether CLARITY captured operative-complexity information beyond both conventional clinical markers and an automated nephrometry-based representation. They were not intended to replace the primary external validation comparisons, which emphasize clinically established and interpretable benchmarks: tumor size and manually assigned R.E.N.A.L. score. Results are shown in Figure **??**.

### Supplementary Figures

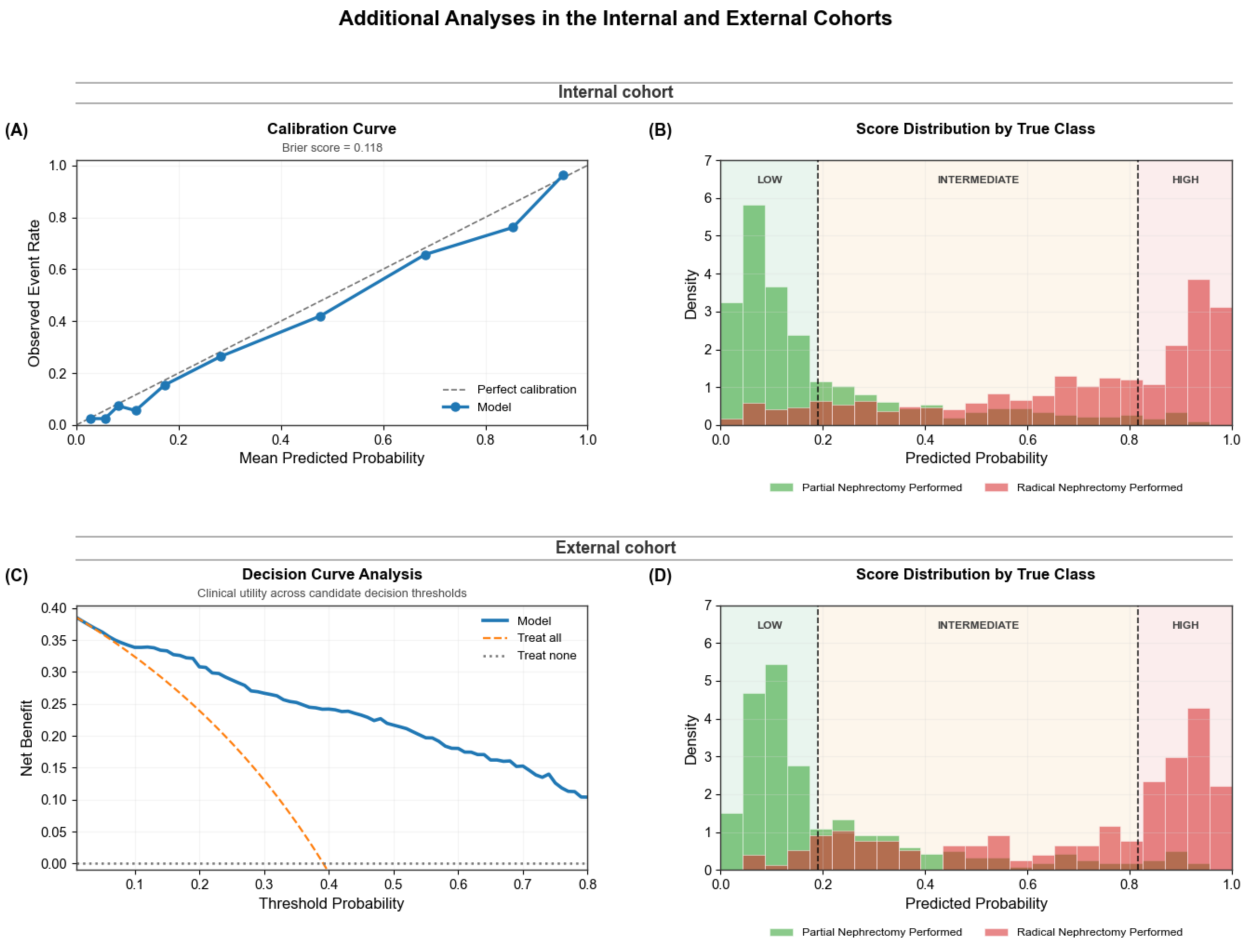


**Figure S1:** Additional internal and external validation analyses. (A) Calibration curve in the internal cohort (Brier score 0.118). (B) Score distribution in the internal cohort. (C) Decision curve analysis in the external cohort. (D) Score distribution in the external cohort.

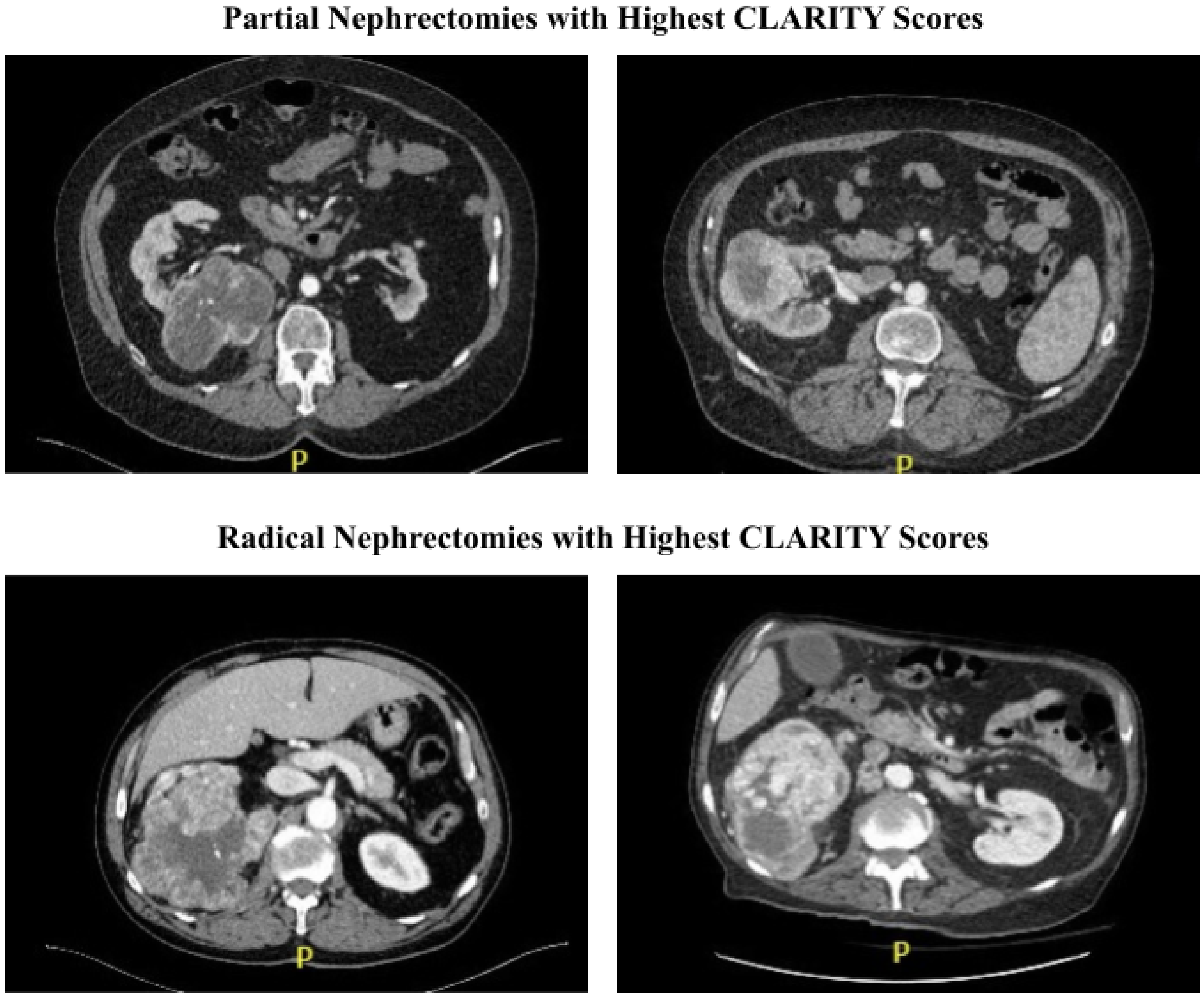


**Figure S2:** Examples of major outliers, including the two patients with partial nephrectomies with the highest CLARITY scores (top) and the two patients with radical nephrectomies with the lowest CLARITY scores (bottom).

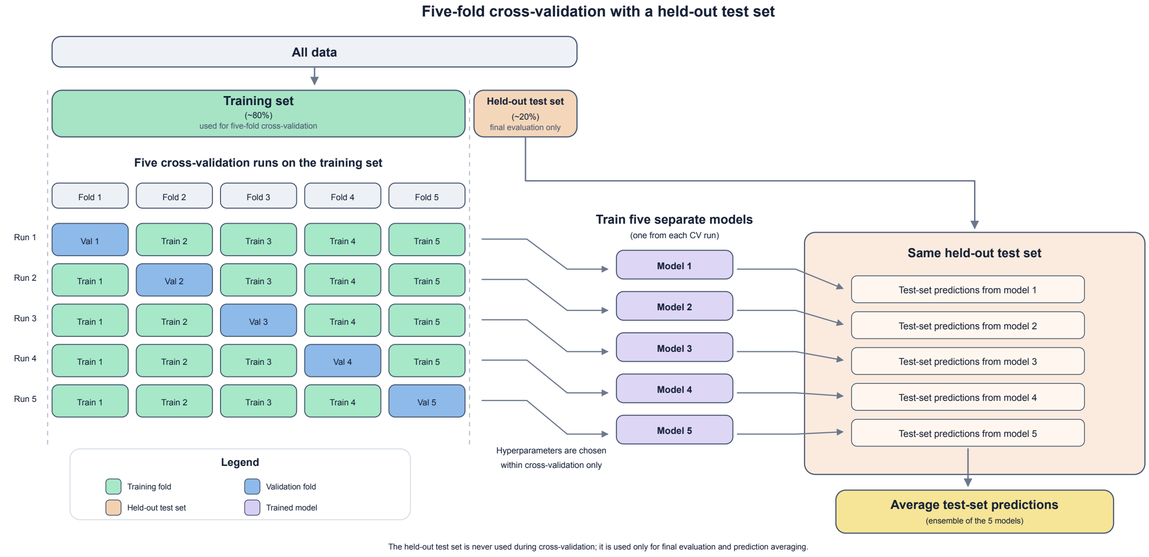


**Figure S3:** Diagram illustrating the overall flow of data in five-fold cross-validation with a held-out test set.

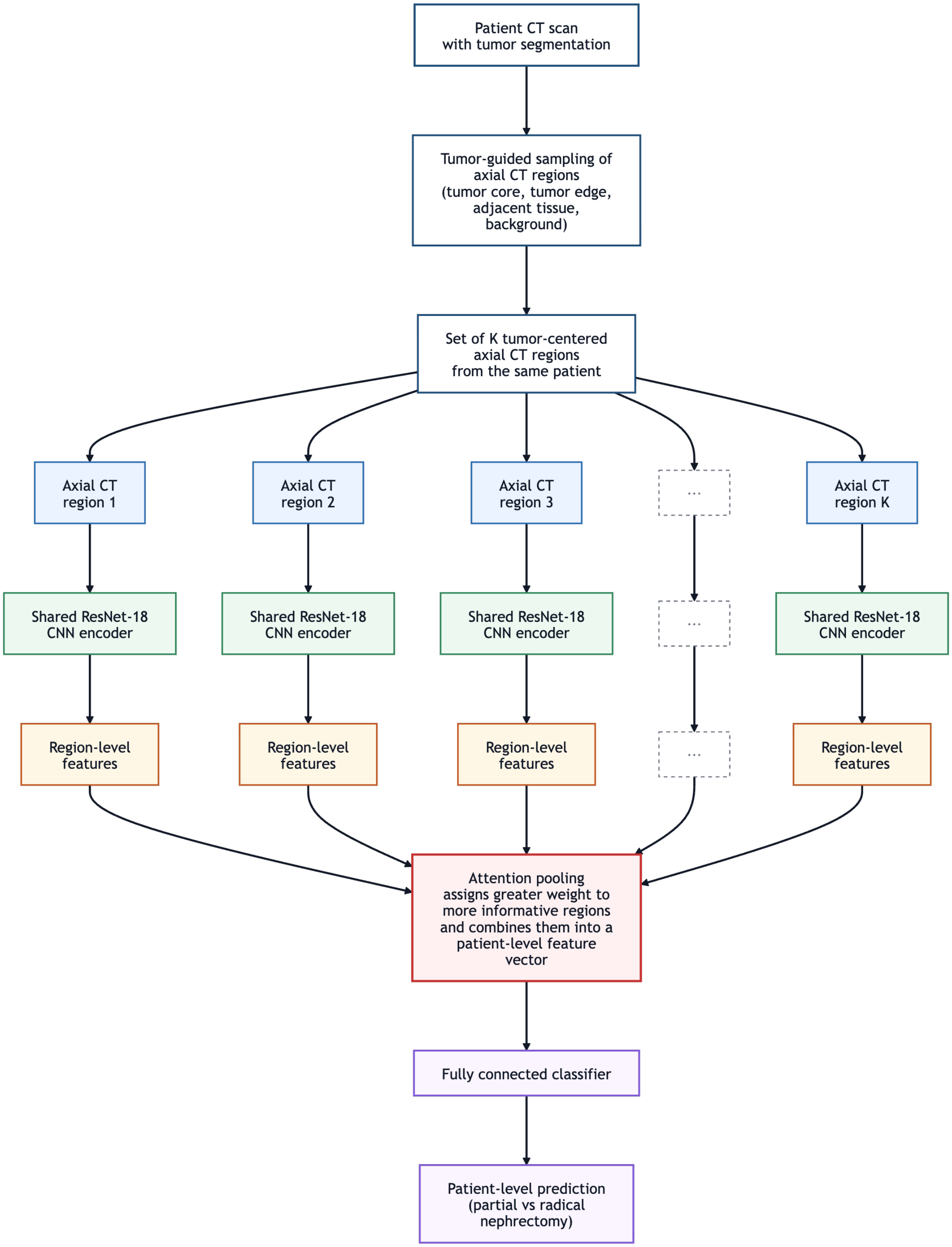


**Figure S4:** Flow chart outlining the model architecture.

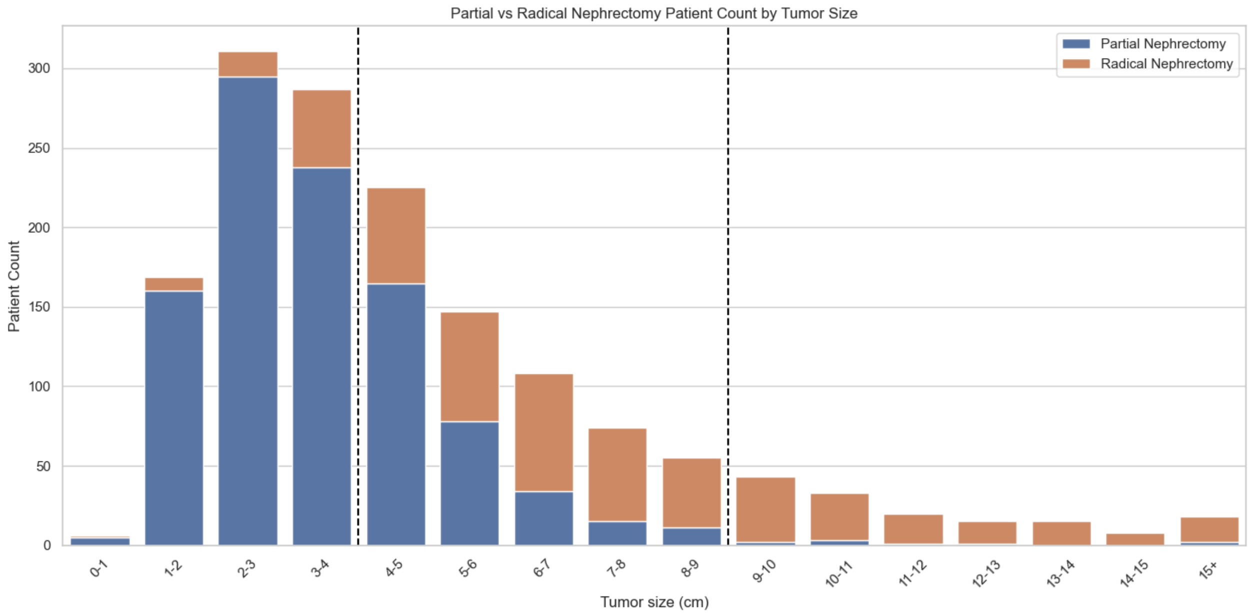


**Figure S5:** The number of patients in the dataset who underwent a partial or radical nephrectomy across tumor size ranges. The vertical dashed lines show the range of the 4–9 cm subset.
